# Development of a Single Molecule Counting Assay to Differentiate Chromophobe Renal Cancer and Oncocytoma in Clinics

**DOI:** 10.1101/2022.05.16.22275137

**Authors:** Khaled Bin Satter, Zach Ramsey, Paul M.H. Tran, Diane Hopkins, Gregory Bearden, Katherine P. Richardson, Martha K. Terris, Natasha M. Savage, Sravan K. Kavuri, Sharad Purohit

## Abstract

Malignant chromophobe renal cancer (chRCC) and benign oncocytoma (RO) are two renal tumor types difficult to differentiate using histology and immunohistochemistry-based methods because of their similarity in appearance. We previously developed a transcriptomics-based classification pipeline with **“**Chromophobe-Oncocytoma Gene Signature” (COGS) on a single-molecule counting platform. Renal cancer patients (n=32, chRCC=17, RO=15) were recruited from Augusta University Medical Center (AUMC). Formalin-fixed paraffin-embedded (FFPE) blocks from their excised tumors were collected. We created a custom single-molecule counting code set for COGS to assay RNA from FFPE blocks. Utilizing hematoxylin-eosin stain, pathologists were able to correctly classify these tumor types (91.8%). Our unsupervised learning with UMAP (accuracy = 0.97) and hierarchical clustering (accuracy = 1.0) identified two clusters congruent with their histology. We next developed and compared four supervised models (random forest, support vector machine, generalized linear model with L2 regularization, and supervised UMAP). Supervised UMAP has shown to classify all the cases correctly (sensitivity = 1, specificity = 1, accuracy = 1) followed by random forest models (sensitivity = 0.84, specificity = 1, accuracy = 1). This pipeline can be used as a clinical tool by pathologists to differentiate chRCC from RO.

**Simple Summary:** We previously reported a gene signature, “Chromophobe-Oncocytoma Gene Signature” (COGS), to differentiate Chromophobe renal cell carcinoma from oncocytoma. Here, we report our results on a single-molecule counting assay with machine learning as a diagnostic pipeline to differentiate chromophobe and oncocytoma tumors utilizing the COGS signature to be used in clinics.

## 1. Introduction

Chromophobe renal cancer (chRCC) and oncocytoma (RO) are renal cancer types, each generally termed as oncocytic renal neoplasms [1] that arise from collecting ducts and constitute about 8-13% of all renal tumors [1,2]. ChRCC is a slow-growing malignant tumor, that once metastasized has a poor prognosis [3]. RO is a benign tumor that does not require treatment [3]. Due to prognostic differences, it is essential to differentiate these tumors to guide treatment. Both of these tumors are diagnosed using hematoxylin and eosin (H&E) and immune-histochemistry (IHC) staining [4]. Histologically these tumors are similar; however, features such as binucleate or multinucleated cells, nuclear wrinkling, perinuclear clearing, presence of mitotic figures, and cytoplasmic invaginations are distinctive to chRCC and can help differentiate chRCC from RO. However, given substantial morphological overlap, distinction between the two may be challenging despite the aforementioned distinctive morphologic characteristics [4]. Due to their similarity in histology and lack of reliable IHC markers, chRCC has a high probability of misdiagnosis as RO.

Immunohistochemical detection of CK7 is used in clinics with variable sensitivity and specificity [4– 6]. Other studies showed that the inclusion of CD110, vimentin, and S100A1 in clinical diagnosis distinguishes chRCC from RO [4,6,7]. Several recent studies [7-9] have suggested using genetic and molecular assays as well as deep learning-based image detection to classify these tumors. Throughout the literature, smaller sample sizes have been the main limitation leading to variation in marker sensitivity and specificity [6,8–10]. In an effort to resolve interobserver variability and misdiagnosis, comprehensive guidelines and additional markers for renal tumor differentiation are needed.

We previously reported the Chromophobe-Oncocytoma-related gene signature (COGS) and a bioinformatics pipeline that can differentiate chRCC from RO and be implemented as a clinical tool [11]. Here we present our results on a retrospective validation of the gene signature at Augusta University Medical Center (AUMC). We developed a diagnostic workflow using a single-molecule counting assay to use formalin-fixed paraffin-embedded (FFPE) samples to differentiate these tumors. We demonstrate the robustness of the gene-signature and bioinformatics pipeline for classification of chRCC from RO with unsupervised and supervised models.

## 2. Materials and Methods

### 2.1 Human subjects and study design

We identified patients (n = 82) who attended AUMC between 2003 and 2018. These patients underwent tumor excision or needle biopsies to diagnose renal masses identified in abdominal images. The inclusion criterion was a histological diagnosis of chRCC or RO. Core/fine needle biopsy samples were excluded. Power analysis showed that six sample per group was required to identify gene expression differences between chRCC and RO for COGS genes (β = 1.65, 1-β = 0.8, α = 0.05). Based on inclusion criteria, we retrieved archived formalin-fixed paraffin-embedded (FFPE) tissues diagnosed as chRCC (n = 17) or RO (n = 15) from the Department of Pathology at AUMC. The tissue samples (n = 32) had been surgically excised from renal tumors and embedded in paraffin after fixing with formalin (FFPE); 5µm thick sections were taken for histological diagnosis (**Figure 1**).

**Figure 1:**
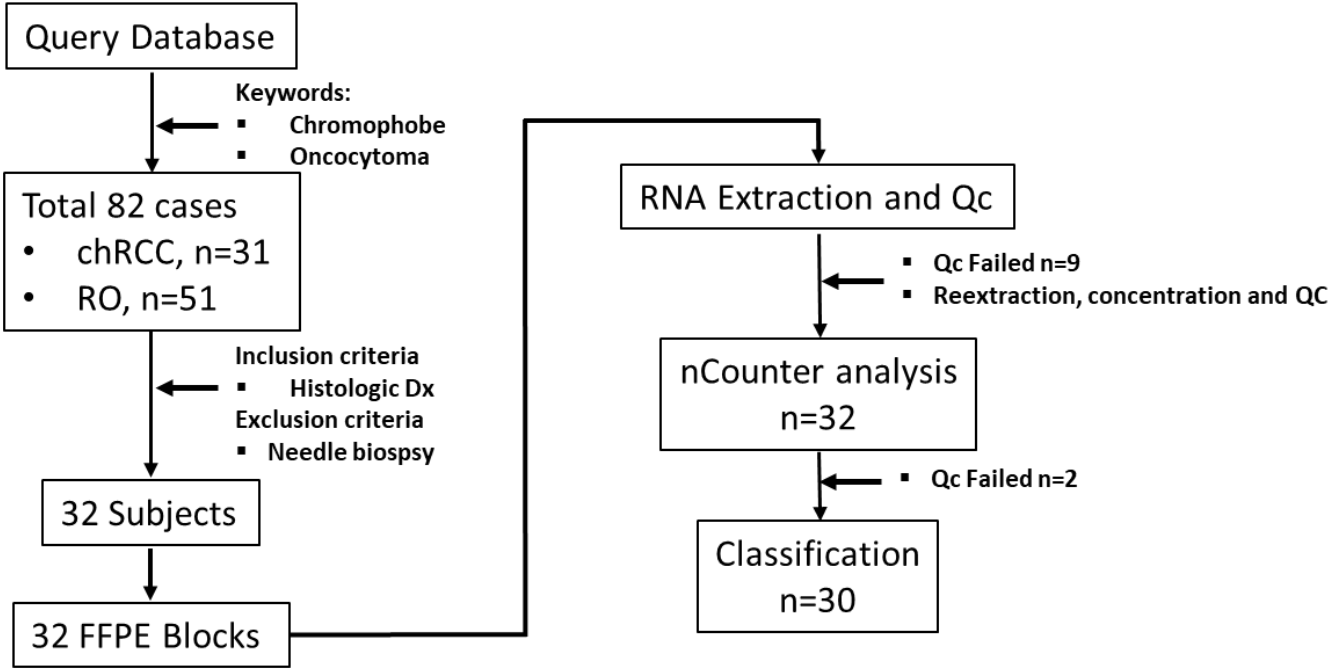
Study workflow from sample collection to RNA quantitation.

Three AUMC Pathologists examined the selected sample set and identified the tumor region on FFPE blocks, while also evaluating the matching H&E slides.

Demographic and clinical data were collected using the Augusta University Cancer Registry and validated through the patients’ electronic health records. The study was conducted according to the Declaration of Helsinki (1997, revised in 2013) and approved by the Institutional Review Board at Augusta University (Biomarkers and Therapeutics in Cancer, 611205-49).

### 2.2 Coring, RNA extraction, and quality control

Tumor-rich region marked by the pathologist was cored using a 2 mm biopsy punch (Integra Miltex, PA, USA). Paraffin was removed from the tissue core by treatment with Citrisolve (Fisher Scientific, Piscataway, NJ, USA) at 65°C. The tissue was then homogenized in 10 mM Tris HCl-EDTA (pH 7.4) by mechanical disruption. According to the manufacturer’s instructions, RNA from lysates was extracted using an RNA extraction kit designed for FFPE tissue (RNEasy, Qiagen, Hilden, Germany). RNA quality, quantity, and concentration were checked using Qbit and TapeStation 2200 (Supplemental Figure 1).

### 2.4 Quantification of gene expression data

Custom nCounter assay consisting of gene expression code set for 33 genes (including three housekeeping genes) was synthesized (NanoString Technology Inc., Seattle, WA, USA) [12]. Two hundred nanograms of RNA/sample were hybridized into the reporter probe. Data capture was performed on a NanoString nCounter Digital Analyzer (NanoString Technology Inc.) and exported as reporter code count (RCC) files. These RCC files were analyzed using nSolver analysis software for quality control purposes (imaging, binding density, positive spike in control, and limit of detection).

The output from nSolver software was read into R for data pre-processing. The data was initially normalized by their concentration, followed by background thresholding. It was normalized first by the geometric mean of the code set’s internal positive controls (Supplemental Figure 2A-B) and then the geometric mean of the housekeeping genes included in our assay (*HPRT1, LDHA*, and *TBP*) (Supplemental Figure 2C-D). The data was then log2-transformed and subjected to our machine learning models described below. We calculated the optimum cut point for each gene by calculating sensitivity plus specificity at each percentile. The value at which sensitivity plus specificity is maximum is called the optimum cut point.

### 2.5 Machine learning models

Unsupervised models were developed using UMAP and hierarchical clustering on NanoString data. Multiple supervised models (random forest, generalized linear model with L2 regularization, and support vector machine) were trained on COGS discovery data [11] with three repeats of 10-fold cross-validation and tested on NanoString data. The discovery data introduced a UMAP manifold for supervised UMAP models and tested on NanoString data. NanoString data’s UMAP projection was used to cluster the data. A contingency matrix was created for all models’ results, and performance metrics (Sensitivity, specificity, positive and negative predictive value) were calculated. The p-value for the metrics was estimated by bootstrap distribution where applicable. Classification metrics, i.e., sensitivity, specificity, positive predictive value, negative predictive value, and accuracy, were calculated by constructing a confusion matrix for each model’s true positive, false positive, true negative, and false negative rate.

### 2.6 Statistical Methods

All statistical analyses were performed using the R language and environment for statistical computing (v4.1.2; R Foundation for Statistical Computing). Software packages were used for cutpointr, caTools, UMAP, ComplexHeatmap, Caret, tidy models in the R environment. Continuous data are presented as mean and standard deviation and differences in the group means were tested using Student’s T-test. Categorical variables are presented as count and percentages and differences in count data were evaluated using Chi-square tests. All p-values were two-sided, and a p < 0.05 was considered significant.

## 3. Results

### 3.1 Human subject demographics

The demographic information on subjects diagnosed with chRCC (n = 17) and RO (n = 15) is presented in Table 1. The mean age of onset of chRCC and RO was 57.8 (SD = 8.51) and 64.3 (SD = 13.9) years. Patients with RO patients significantly are older than chRCC (p-value = 0.003). There is a higher number of females in RO, overall, there were no significant differences in distribution of male and female in both tumor types (Table 1). All chRCC tumors were staged. The majority of the cases were Stage I (64%), whereas Stage II and III were 17% each. RO cases were not staged. The race breakdown for chRCC was 70% Caucasian and 29% African American, while RO was 60% Caucasian and 33% African American.

**Table 1:** Clinical and Demographic information for chRCC and RO cases in the AUMC cohort.

| Variable | chRCC<sup>a</sup> | RO<sup>b</sup> | Overall | p-value |
| --- | --- | --- | --- | --- |
| Stage, n(%) |  |  |  |  |
| 1 | 11(64.7%) | NA | 11(34.4%) |  |
| 2 | 3(17.6%) | NA | 3(9.4%) |  |
| 3 | 3(17.6%) | NA | 3(9.4%) |  |
| Age at Diagnosis, Mean (SD) | 57.8(14.7) | 71.5(8.51) | 64.3(13.9) | 0.003* |
| Race, n (%) |  |  |  |  |
| AA | 5(29.4%) | 5(33.3%) | 10(31.3%) | 0.55** |
| C | 12(70.6%) | 9(60%) | 21(65.6%) |  |
| Other | 0 | 1(6.7%) | 1(3.1%) |  |
| Gender, n (%) |  |  |  |  |
| F | 8(47.1%) | 3(20%) | 11(34.4%) | 0.11** |
| M | 9(52.9%) | 12(80%) | 21(65.6) |  |

<sup>a</sup>chRCC: chromophobe renal cell carcinoma, <sup>b</sup>RO: renal oncocytoma cases are not staged, \*Student's T-test, \*\*Chi-Square test

### 3.3 Univariate analysis

We performed univariate AUC analysis for differentiating chRCC and RO with count difference. The sensitivity was 1 for nine genes (*AQP6, NDUFS1, MAP4K3, HOOK2, ESRP1, ELMO3, BSPRY, PRDX3, and LIMS1*) with a median of 0.94. Seven genes have a specificity of 1 (*AQP6, AP1M2, ITGB3, LRFN5, RSPO3, SPINT2, and LSR*), and a median of 0.93. The minimum AUC was 0.5 (*LAMA1*) with a median of

0.93 and a maximum of 1 (*AQP6)*. Genes with the highest difference in AUC analysis were *AQP6* (AUC = 1.00), *BSPRY* (AUC = 0.99), *HOOK2* (AUC = 0.99), *SPINT2* (AUC = 0.99), *ESPR1* (AUC = 0.99), *ITGB3* (AUC = 0.98). The maximum is 1 (*AQP6*) for accuracy, and the median is 0.90. Amongst all the COGS genes, *AQP6* has the highest sensitivity, specificity, AUC, and accuracy. The full table is presented in Table 2.

**Table 2:** Univariate analysis of chRCC and RO NanoString Data. All values were calculated at the optimum cut point between chRCC and RO count data

| Gene | Optimal cut point | Accuracy | AUC | Sensitivity | Specificity |
| --- | --- | --- | --- | --- | --- |
| *AP1M2* | 10.16 | 0.97 | 0.96 | 0.94 | 1 |
| *AQP6* | 13.24 | 1 | 1 | 1 | 1 |
| *ATP2C1* | 11.38 | 0.87 | 0.94 | 0.94 | 0.79 |
| *BSPRY* | 9.2 | 0.97 | 0.99 | 1 | 0.93 |
| *CLDN8* | 12.57 | 0.93 | 0.93 | 0.94 | 0.93 |
| *DNAI3* | 6.51 | 0.9 | 0.93 | 0.86 | 0.94 |
| *ELMO3* | 9.32 | 0.97 | 0.97 | 1 | 0.93 |
| *ESRP1* | 9.94 | 0.97 | 0.99 | 1 | 0.93 |
| *HOOK2* | 9.02 | 0.97 | 0.99 | 1 | 0.93 |
| *ITGB3* | 9.97 | 0.97 | 0.98 | 0.93 | 1 |
| *KCNG3* | 7.62 | 0.8 | 0.83 | 0.86 | 0.75 |
| *KIDINS220* | 9.8 | 0.9 | 0.92 | 0.93 | 0.88 |
| *KRT7* | 8.28 | 0.9 | 0.96 | 0.94 | 0.86 |
| *LAMA1* | 5.63 | 0.6 | 0.5 | 0.36 | 0.81 |
| *LIMS1* | 9.26 | 0.97 | 0.98 | 1 | 0.94 |
| *LRFN5* | 9.55 | 0.8 | 0.88 | 0.63 | 1 |
| *LSR* | 10.43 | 0.93 | 0.96 | 0.88 | 1 |
| *MANEA* | 8.66 | 0.9 | 0.9 | 0.86 | 0.94 |
| *MAP4K3* | 8.62 | 0.93 | 0.98 | 1 | 0.88 |
| *MSH2* | 8.37 | 0.67 | 0.54 | 0.86 | 0.5 |
| *NDUFS1* | 11.33 | 0.9 | 0.91 | 1 | 0.81 |
| *PLCL1* | 10.78 | 0.87 | 0.9 | 0.93 | 0.81 |
| *PLCL2* | 11.18 | 0.73 | 0.75 | 0.56 | 0.93 |
| *PNPT1* | 8.68 | 0.87 | 0.82 | 0.86 | 0.88 |
| *PRDX3* | 11.94 | 0.97 | 0.96 | 1 | 0.94 |
| *RSPO3* | 8.83 | 0.83 | 0.88 | 0.69 | 1 |
| *S100A1* | 7.12 | 0.87 | 0.88 | 0.93 | 0.81 |
| *SOCS1* | 9.32 | 0.93 | 0.98 | 0.94 | 0.93 |
| *SPINT2* | 13 | 0.97 | 0.99 | 0.94 | 1 |
| *SUCLA2* | 10.83 | 0.87 | 0.85 | 0.86 | 0.88 |

AUC: Area under the curve.

### 3.3 Unsupervised and supervised machine learning models

As single markers can fail to differentiate chRCC and RO, we implemented multivariate models using unsupervised learning. We implemented UMAP and hierarchical clustering to see the combined markers’ ability to differentiate these tumors. UMAP analysis on NanoString count data showed two clusters, and 29/30 samples were correctly identified (Supplemental Figure 3). Then we developed a supervised UMAP model using COGS discovery data as the training set by setting parameters to n_neighbors = 15, epoch = 200, min_dist = 0.1, init = “spectral”, metric = “Euclidean” (Figure 2A) [11]. This trained model was tested on AUMC cohort. This trained UMAP model created two clusters congruent with their histology (30/30 samples are correctly classified) (Figure 2B). Hierarchical clustering showed 100% congruency with their histological classification (Figure 2C).

**Figure 2:**
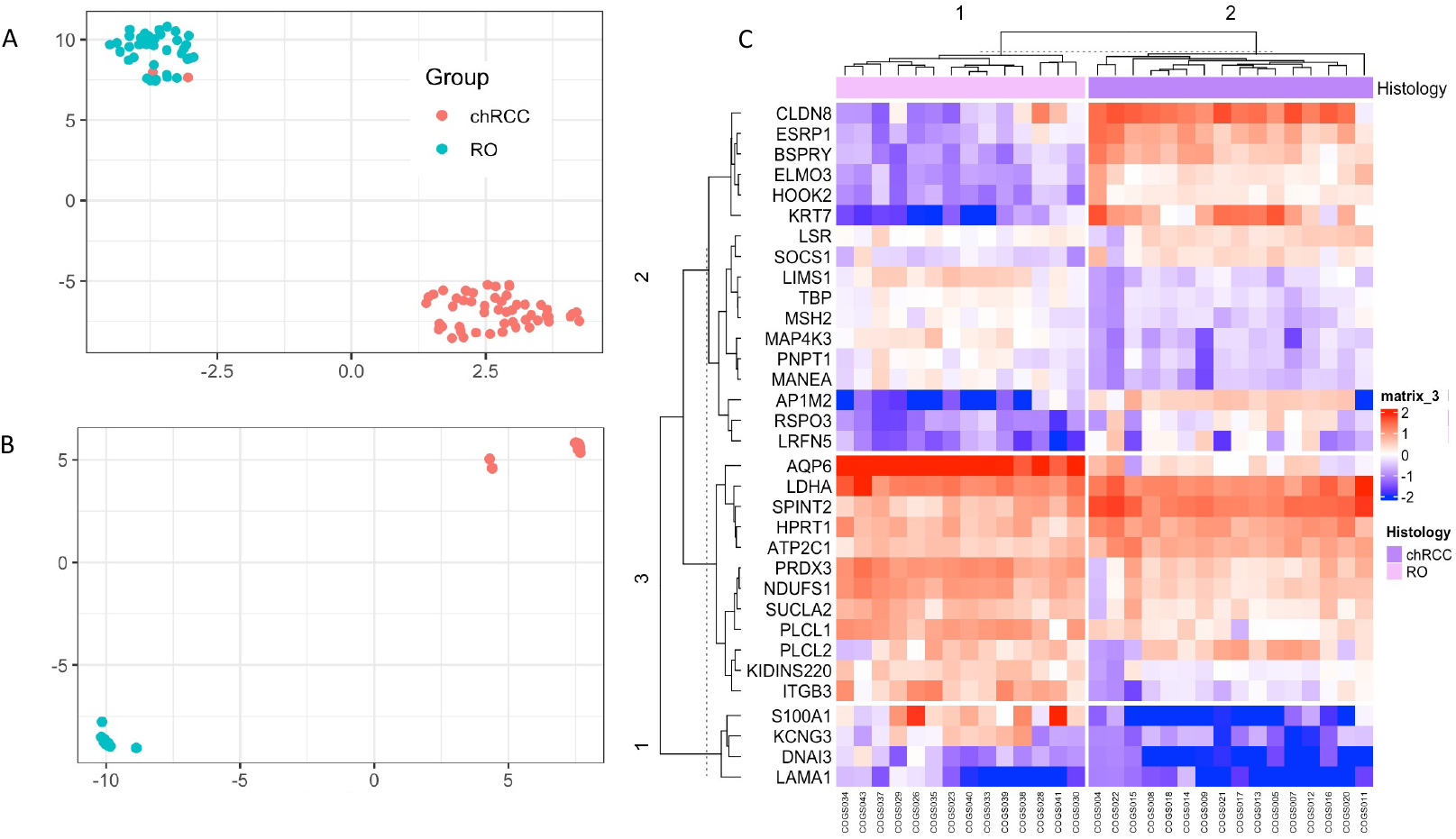
Unsupervised classification of the NanoString data with UMAP and Hierarchical Clustering. A: UMAP of discovery data showing two clusters were used to train the supervised UMAP model. B: UMAP projection of the NanoString data using the trained UMAP model from A. C: Hierarchical clustering showing two clusters for the samples, congruent with their histological classification.

Although unsupervised models are a powerful tool for identifying data structure in high dimensional space, they are not ideal for testing a new sample. Therefore, we compared four supervised learning models viz., random forest, support vector machine, generalized linear model with L2 regularization trained with COGS’ discovery data with 10-fold cross-validation with three repeats. The four models were compared (Table 3) by sensitivity, specificity, positive predictive value, negative predictive value, and accuracy.

**Table 3:** Supervised Model metrics for classification of chRCC and RO.

| Metric | Random Forest | SVM | GLM | Supervised UMAP |
| --- | --- | --- | --- | --- |
| Sensitivity | 0.84 | 0.76 | 0.88 | 1 |
| Specificity | 1 | 1 | 1 | 1 |
| Accuracy | 0.93 | 0.83 | 0.9 | 1 |
| 95% CI | 0.78-0.99 | 0.65-0.94 | 0.73-0.98 | - |
| p-value | $4.74 \times 10^{-5}$ | 0.07659 | 0.001066 | ∞ |
| PPV | 1 | 1 | 1 | 1 |
| NPV | 0.87 | 0.64 | 0.78 | 1 |

SVM: Support Vector Machine, GLM: Generalized Linear Model, UMAP: Uniform Manifold Approximation and Projection, PPV: Positive Predictive Value, NPV: Negative Predictive Value

Supervised UMAP showed to be most accurate in classifying the chRCC and RO based on their COGS expression (Sensitivity = 1.0, Specificity = 1.0, PPV = 1.0, NPV = 1.0 and Accuracy = 1.0). As Supervised UMAP did not have any misclassification, the p-value is ∞;, and the 95% confidence interval cannot be calculated. Random forest models performed better than the other SVM and GLMNet models. For random forest models, a minimum of 250 trees were needed to achieve an accuracy of 1 (Figure 3A). A sample tree is presented in Figure 3B. Although support vector machines are popular in classification analysis, they did not reach statistical significance in our dataset (Figure 3C, Table 3). Coefficients for the generalized linear model with ridge regression are presented in Figure 3D. All the supervised models were used to create a confusion matrix, and sensitivity, specificity, positive predictive value, negative predictive value, and accuracy were calculated (Table 3). All the p-values were calculated by accuracy over no information rate for the models.

**Figure 3:**
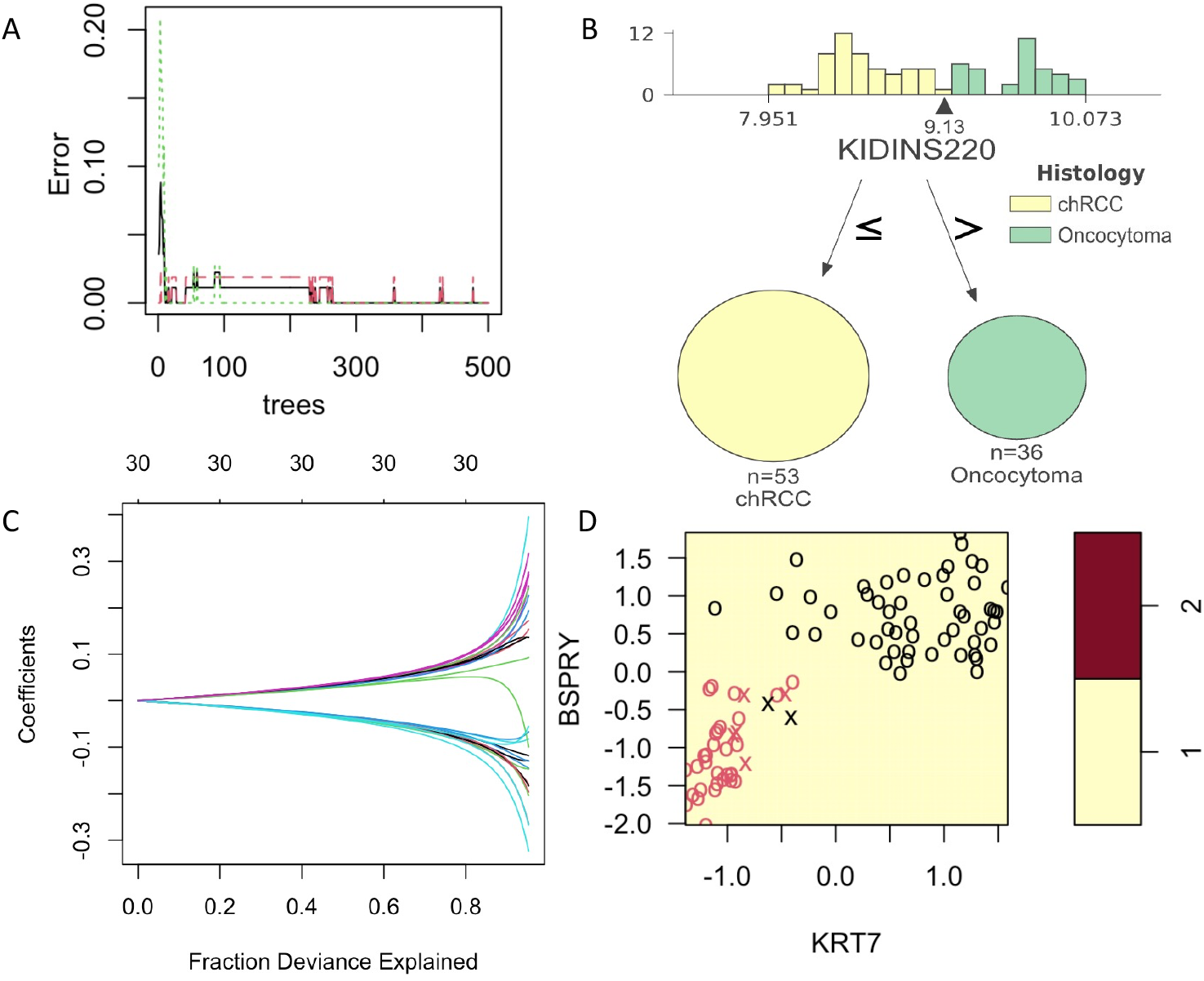
Supervised models with training on COGS discovery data and tested on NanoString data. A: Figure 3: Development and comparison of four supervised models. Random Forest models showing a minimum of 250 trees can achieve the error = 0. B: A sample tree showing the distribution of gene *KIDINS220* expression on COGS discovery data with the arrow pointing at the best cutoff value. C: A sample support vector model with *KRT7* and *BSPRY*. D: Generalized linear model with Ridge regression showing the fraction explained by the COGS genes as covariates. All these metrics show the results on NanoString data (AUMC cohort).

### 3.4 H&E scoring between pathologists

Three independent AUMC pathologists evaluated H&E for the selected sample set (n = 32). Two of the three pathologists had one discrepancy in diagnosis from the original clinical diagnosis, while the third pathologist had two discrepancies. We developed a linear mixed model with clinical diagnosis as the fixed effects and H&E and scorer as the random effects. The model showed that H&E accounts for 0.918 (p-value <0.001) of the variation in the diagnosis of these cases. The student t-test between the clinical diagnosis and pathologist impression or between the pathologist impression did not show any difference (p-value > 0.05). All the raw data is provided in Supplemental Table 1.

## 4. Discussion

In this study, we tested the potential of COGS to differentiate chRCC and RO renal tumor types. We also developed a single-molecule counting assay in a single institutional cohort. Our univariate analysis demonstrates that the majority of the genes in the COGS signature shows expression level difference between the tumor types and can serve as a potential biomarker to differentiate these tumors (Table 2). We showed that the COGS profile between these tumors is distinct in unsupervised models (UMAP and hierarchical clustering) (Figure 2B-C). To classify future samples, we developed and compared four different supervised models (random forest, support vector machine, generalized linear model, and supervised UMAP) (Figure 3A-D) and identified supervised UMAP outperforming all the other models in classifying chRCC and ROs (Table 3). This workflow can be implemented in clinical settings to quantify and correctly classify future samples suspected of chRCC or RO.

The current clinical workflow to differentiate these tumors is based on H&E impressions and certain IHC markers. H&E with or without IHC suffers in sensitivity and specificity because there is a lacking of definitive features on H&E or the overlapping expression of the immunohistochemical markers. This is even more difficult in biopsy cases where there is limited tissue. CK7 is the most widely used marker specific for chRCC with variable sensitivity (0.73-1.0) and specificity (0.84-1.0) [13,14]. S100A1 is the specific IHC marker for RO with variable sensitivity (0.87-1.0) and specificity (0.7-1.0) [15,16]. The variability in these markers leaves ∼10% of cases to be misdiagnosed by traditional clinical workflow. IHC staining can be subjective and requires optimization as the reading is dependent on the quality of the staining [17]. In comparison, gene expression workflows follow a strict protocol and have high quality-control measures. Our NanoString validation pipeline correctly identified 100% of the cases analyzed in this study. The clinical workflow can overcome the obstacles such as interobserver variability, with the integration of supervised learning. *AQP6, HOOK2, AP1M2, ESPR1*, and *CLDN* are from the COGS panel in the previous study [18,19]. We report *AQP6, AP1M2, LIMS1, PRDX3, SPINT2, BSPRY, ELMO3, ESPR1, HOOK2, ITGB3 (*sensitivity > 0.93, specificity > 0.93, AUC > 0.96 and accuracy > 0.93) as top-performing genes in our study and can serve as potential transcriptomic biomarkers between these tumors.

A limitation of using bulk transcriptomics is that the expression profile includes tumor cells and stromal cells, immune cells, etc. Future studies are needed to identify the differences between tumor cells themselves in chRCC and RO. Patients with RO were older in age compared to chRCC as peak age of detection of RO is seventh decade whereas peak chRCC detection is sixth decade [20]. Another limitation was due to the FFPE RNA being highly fragmented, which resulted in some genes not showing the observed differences in our discovery data, such as LAMA1 and MSH2. To work around the fragmentation issues, multivariate models were implemented as they account for such variation due to technical or biological reasons.

The strength of this study is the direct clinical application. The supervised model training was on bulk transcriptomic data from multiple GEO studies previously validated in microarray and RNASeq platform, which are now translated into the NanoString platform. Therefore, to quantify the expression of COGS genes, assayed on any platform can be conducted and followed up by the implementation of the machine learning models. This workflow can be performed directly on biopsies by extracting RNA and quantifying it using the NanoString platform. Our method has shown an overall greater accuracy (Supervised UMAP, accuracy = 1.0) in comparison to the current clinical workflow [13,15,16,21].

We have shown that the single-molecule counting on the nCounter platform working in conjunction with our machine-learning pipeline can be a viable clinical assay for differentiation of chRCC from RO. This approach has been shown to predict therapeutic outcomes in the breast cancer [22], diagnosis of small B-cell lymphoid neoplasm [23] cancer, and classification of gliomas [24].

## 5. Conclusions

We report a pipeline that uses gene expression profiling by single molecule counting to differentiate chRCC from RO, which can be an important clinical tool in renal cancer diagnosis.

## Supporting information

Suppementary material

## Data Availability

The data and code presented in this study are available on GitHub.

https://github.com/kbsatter/chRCC-paper-2

## Author Contributions

“Conceptualization, KBS, PMHT, MKT; methodology, KBS, ZR, PMHT, SP.; formal analysis, KBS, PMHT, SP.; investigation, KBS, ZR, PMHT, NMS, SKK, SP; resources, ZR, DH, GB, NMS, SKK, SP; data curation, KBS, PMHT; writing—original draft preparation, KBS.; writing—review and editing, ZR, PMHT, KPR, NMS, SKK, SP; visualization, KBS, PMHT, SP; supervision, NMS, SKK, SP; project administration, DH, GB, SP; funding acquisition, DH, GB, SP. All authors have read and agreed to the published version of the manuscript.

## Funding

This work is supported by an institutional grant for the Biomarkers and Therapeutics for Cancers (BAT) study. PMHT was supported by the NIH/NIDDK fellowship (F30DK121461). Institutional funds also provided Article-processing charges.

## Institutional Review Board Statement

The study was conducted according to the guidelines of the Declaration of Helsinki and approved by the Institutional Review Board of Augusta University (protocol code 611205-37 and approved on 3 April 2020)

## Informed Consent Statement

The tissue blocks and the phenotype data collected were de-identified. Therefore, the patient consent was waived by the IRB.

## Data Availability Statement

The data and code presented in this study are available on GitHub (https://github.com/kbsatter/chRCC-paper-2).

## Acknowledgments

The authors acknowledge the support of the Pathology staff at the Department of Pathology, Medical College of Georgia, Augusta University.

## Conflicts of Interest

The authors declare no conflict of interest.

## Notes

### Competing Interest Statement

The authors have declared no competing interest.

### Author Declarations

: The study was conducted according to the guidelines of the Declaration of Helsinki and ap-proved by the Institutional Review Board of Augusta University (protocol code 611205-37 and approved on 3 April 2020)

