## Supplementary material for "Development of a Single Molecule Counting Assay to Differentiate Chromophobe Renal Cancer and Oncocytoma in Clinics": Suppementary material

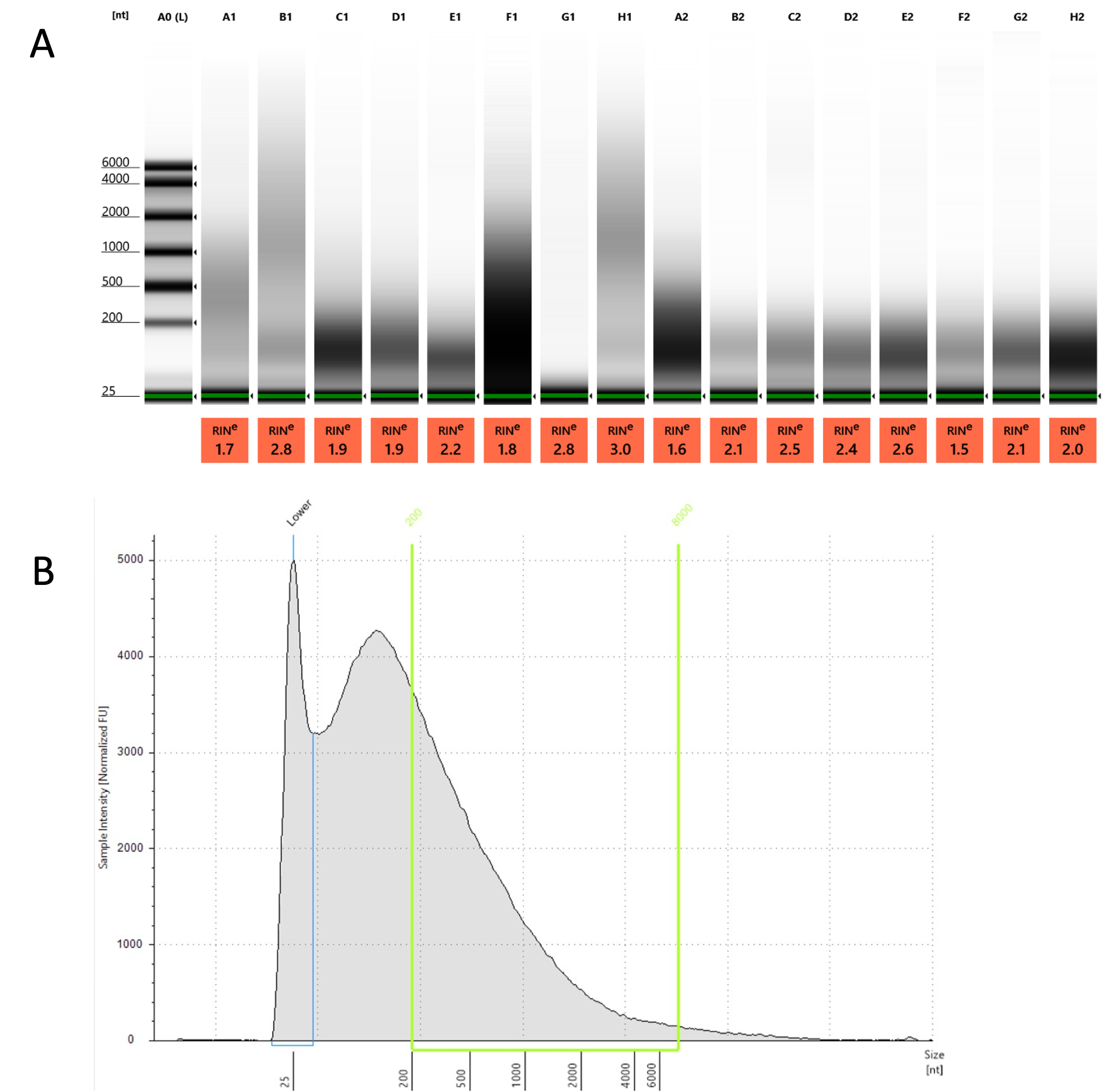

Supplemental Figure 1: RNA quality control on Tapestation 2200. A: Gel images showing RNA quality. The bottom number represents RIN (RNA integrity number). B: Showing a representative sample for RNA fragments. The green horizontal line represents the target range for hybridization. The RNA concentration was calculated for this range for each sample.

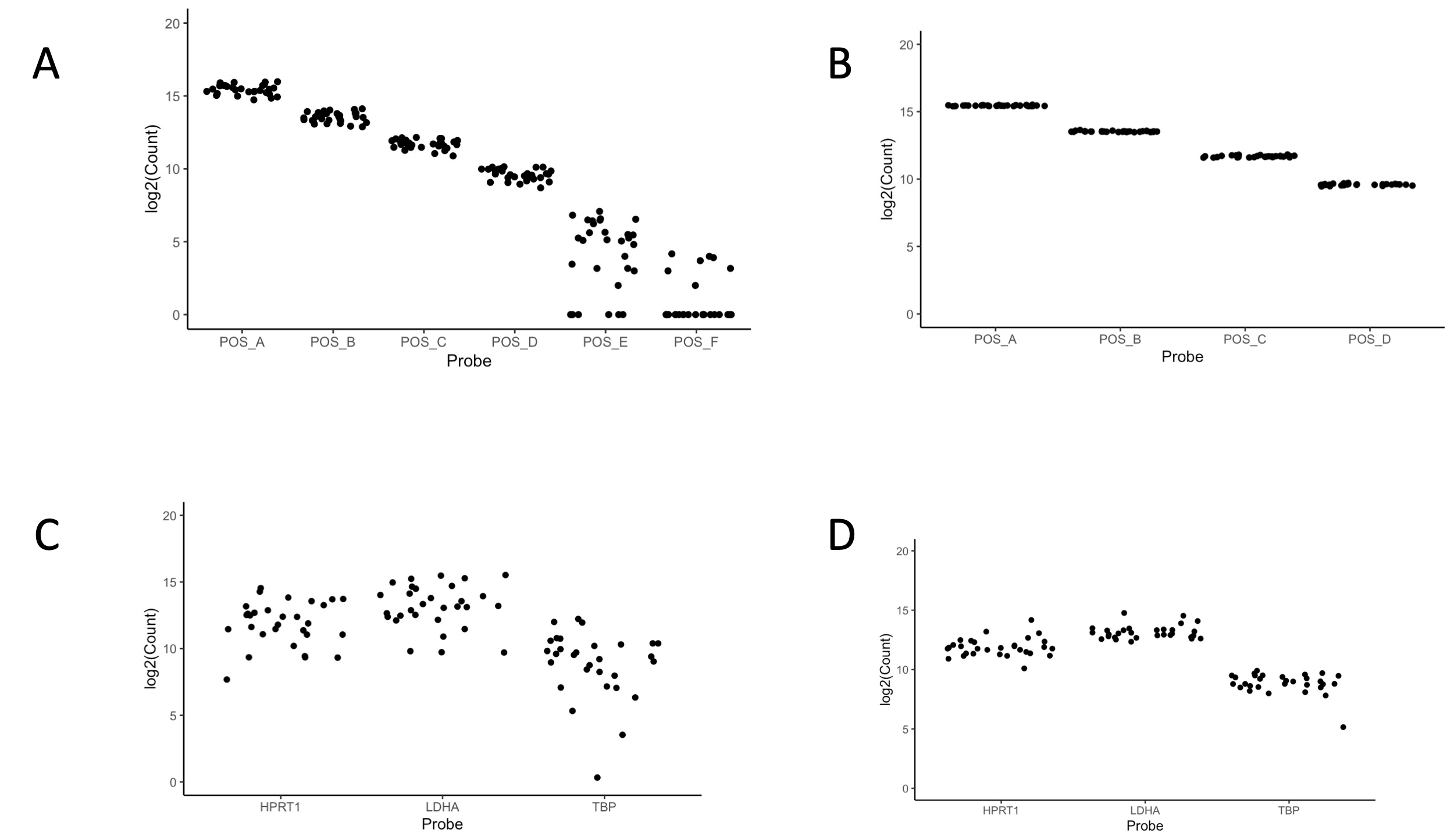

Supplemental Figure 2: Positive control and housekeeping gene normalization using geometric mean. A: Before normalization of the positive control B: After positive control normalization. C: Before housekeeping gene normalization. D: After housekeeping gene normalization

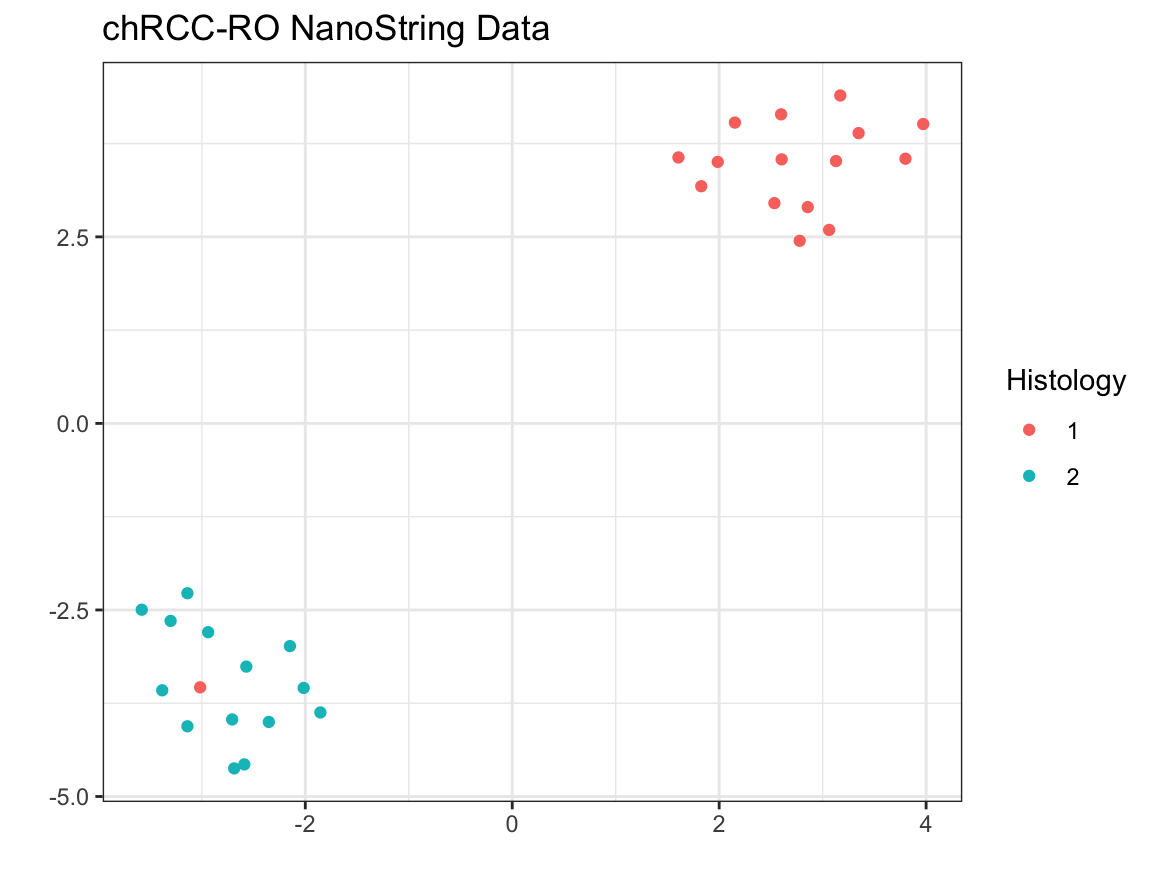

Supplemental Figure 3: Uniform manifold approximation and projection (UMAP) analysis showing two clusters. Raw count data was normalized with internal controls and log2 transformed prior to UMAP analysis. Histology 1 represents chRCC and 2 represents RO samples.

Supplemental Table 1: Comparison of original hematoxyl-eosin stained sections at the time of diagnosis with re-review of the slides by three independent pathologists. The identity and diagnosis was blinded to be reviewed by pathologists.

| Sample ID | Histology | Reviewer 1 | Reviewer 2 | Reviewer 3 |
| --- | --- | --- | --- | --- |
| Rand1 | chRCC | chRCC | chRCC | chRCC |
| Rand10 | chRCC | chRCC | chRCC | chRCC |
| Rand11 | chRCC | chRCC | chRCC | chRCC |
| Rand12 | RO | RO | RO | RO |
| Rand13 | chRCC | chRCC | chRCC | chRCC |
| Rand14 | chRCC | chRCC | chRCC | chRCC |
| Rand15 | RO | RO | RO | RO |
| Rand16 | chRCC | chRCC | chRCC | chRCC |
| Rand17 | RO | RO | RO | RO |
| Rand18 | chRCC | chRCC | chRCC | chRCC |
| Rand19 | RO | RO | RO | RO |
| Rand2 | chRCC | chRCC | chRCC | chRCC |
| Rand20 | chRCC | chRCC | chRCC | chRCC |
| Rand21 | RO | RO | RO | RO |
| Rand22 | RO | RO | RO | RO |
| Rand23 | RO | RO | RO | RO |
| Rand24 | RO | RO | RO | RO |
| Rand25 | chRCC | chRCC | chRCC | chRCC |
| Rand26 | chRCC | chRCC | chRCC | chRCC |
| Rand27 | chRCC | chRCC | chRCC | chRCC |
| Rand28 | RO | RO | RO | RO |
| Rand29 | chRCC | chRCC | chRCC | chRCC |
| Rand3 | chRCC | chRCC | chRCC | RO |
| Rand30 | chRCC | chRCC | chRCC | chRCC |
| Rand31 | chRCC | RO | RO | RO |
| Rand32 | RO | RO | RO | RO |
| Rand4 | chRCC | chRCC | chRCC | chRCC |
| Rand5 | RO | RO | RO | RO |
| Rand6 | RO | RO | RO | RO |
| Rand7 | RO | RO | RO | RO |
| Rand8 | RO | RO | RO | RO |
| Rand9 | RO | RO | RO | RO |

Software and packages:

All statistical analyses were performed using the R language and environment for statistical computing (v4.1.2; R Foundation for Statistical Computing).

1. Quality control: NanoString nCounter output (RCC – reporter code count) was read by nSolver [1] for quality control for imaging QC, binding density, positive spike control and limit of detection QCs.
2. Determination of cutpoint: Optimum cut points were determined by cutpointr [2].
3. AUC analysis: AUC values between the tumor types were analyzed by catools [3].
4. Unsupervised learning: Unsupervised learning was implemented from UMAP [4] and Hierarchical clustering by ComplexHeatmap [5]. UMAP components were used as “features”. Hierarchical Clustering was implemented from Complex heatmap (clustering distance = “maximum” and clustering method = “ward.D”).
5. Supervised learning: Supervised learning models were Random forest, support vector machine and generalized linear model implemented from Caret [6] package. Supervised UMAP was implemented from tidymodels [7].
6. Linear Mixed model: Linear mixed models were implemented from lme4 [8].

All codes and data are available at GitHub (<https://github.com/kbsatter/chRCC-paper-2> ).

1.	NSolver Advanced Analysis Software Available online: https://nanostring.com/products/analysis-solutions/nsolver-advanced-analysis-software/ (accessed on 9 May 2022).

2.	Thiele, C. *Cutpointr: Determine and Evaluate Optimal Cutpoints in Binary Classification Tasks*; 2021;

3.	Tuszynski, J. *CaTools: Tools: Moving Window Statistics, GIF, Base64, ROC AUC, Etc*; 2021;

4.	McInnes, L.; Healy, J.; Melville, J. UMAP: Uniform Manifold Approximation and Projection for Dimension Reduction. *arXiv:1802.03426 [cs, stat]* **2020**.

5.	Gu, Z. *ComplexHeatmap: Make Complex Heatmaps*; Bioconductor version: Release (3.12), 2021;

6.	Kuhn, M.; Wing, J.; Weston, S.; Williams, A.; Keefer, C.; Engelhardt, A.; Cooper, T.; Mayer, Z.; Kenkel, B.; R Core Team; et al. *Caret: Classification and Regression Training*; 2020;

7.	Tidymodels Available online: https://www.tidymodels.org/ (accessed on 29 April 2022).

8.	Bates, D.; Mächler, M.; Bolker, B.; Walker, S. Fitting Linear Mixed-Effects Models Using Lme4. *Journal of Statistical Software* **2015**, *67*, 1–48, doi:10.18637/jss.v067.i01.
